# Estimation of the Lifetime Quality-Adjusted Life Years (QALYs) Lost Due to Syphilis acquired in the United States in 2018

**DOI:** 10.1101/2022.01.26.22269934

**Authors:** Kyueun Lee, Shiying You, Yunfei Li, Harrell Chesson, Thomas L. Gift, Andrés A. Berruti, Katherine Hsu, Reza Yaesoubi, Joshua A. Salomon, Minttu Rönn

## Abstract

**Background:** The purpose of this study was to estimate the health impact of syphilis in the United States in terms of the number of quality-adjusted life years (QALYs) lost attributable to infections in 2018.

**Methods:** We developed a Markov model which simulates the natural history and long-term sequelae of syphilis. The model was parameterized by sex (men and women), sexual orientation (women who have sex with men, men who have sex with women [MSW], and men who have sex with men [MSM]), and by age at primary infection. We developed a separate decision tree model to account for health losses due to congenital syphilis. We estimated the average lifetime number of QALYs lost per infection, and the total expected lifetime number of QALYs lost due to syphilis acquired in 2018. We performed probabilistic sensitivity analysis to account for uncertainty in the model’s estimates.

**Findings:** We estimated the average number of discounted lifetime QALYs lost per infection as 0.09 [0.03-0.19 95% uncertainty interval (UI)]. The QALY loss per infection was lower among MSM (0.06) than among MSW (0.15) and women (0.10). The total expected number of QALYs lost due to syphilis acquired in 2018 was 13,349 (5,071-31,360 95%UI). MSM account for 6,373 (47.7%) of the overall burden, compared to MSW (32.1%) and women (20.2%). For each case of congenital syphilis, we estimated 1.79 (1.43-2.16 95%UI) QALYs lost for the child and 0.06 (0.01-0.14 95%UI) QALYs lost for the mother. These per-case estimates correspond to 2,332 (1,871-2,825 95%UI) and 79 (17-177 95%UI) QALYs lost for children and mothers, respectively, due to congenital syphilis in 2018.

**Conclusion:** Syphilis causes substantial health losses in adults and children. Quantifying these health losses in terms of QALYs can inform cost-effectiveness analyses and can facilitate comparisons of the burden of syphilis to that of other diseases.

## Introduction

Syphilis, a sexually transmitted genital ulcerative disease caused by infection with *Treponema pallidum*, can cause a range of adverse health outcomes if left untreated, including severe outcomes relating to pregnancy. In the United States in 2018, there were 115,045 diagnosed syphilis cases, the highest number reported since 1991, and 1,306 reported cases of congenital syphilis, which was threefold higher than diagnoses in 2013 (1). In light of this large and rising burden, estimates of the lifetime costs and number of quality-adjusted life years (QALYs) lost per infection can quantify the public health and economic impact of syphilis infections and inform cost-effectiveness analyses of interventions to prevent syphilis. A recent modelling study on the economic impact of syphilis in the United States estimated that the average discounted lifetime cost per infection is $1190 (2). However, to our knowledge, no published studies have assessed the average number of QALYs lost per infection or the overall annual QALY losses attributable to syphilis.

Measuring the health impact of syphilis in adults and congenital syphilis using a standard health metric like QALYs has a benefit of combining the morbidity and mortality impact into one measure(3). The QALY is a measure of disease burden which incorporates morbidity and mortality. Health conditions like syphilis can cause reductions in the quality and length of life, thereby reducing the number of QALYs that would have been achieved in the absence of syphilis. One QALY represents one year in perfect health. Assessing the burden of syphilis in terms of QALYs also allows for comparing the burden of syphilis with that of other health outcomes.

The purpose of our study was to estimate the average number of QALYs lost per infection and the total number of QALYs lost due to syphilis acquired in the United States in 2018 by using a Markov model that simulates long-term sequelae of syphilis infection.

## Methods

### Model overview

Our Markov model consists of two parts: the natural history that describes disease progression and testing and treatment that prevents further disease progression. For syphilis natural history, we adopted the framework used in Tuite and colleagues (4). The model is run by sex (men and women), sexual orientation (women who have sex with men, men who have sex with women, and men who have sex with men), and by age at primary infection. The model calculates the lifetime number of QALYs lost per infection by assessing the disutility associated with syphilis, in monthly intervals. We estimated average lifetime QALYs lost per infection in three subpopulations: men who have sex with men (MSM), men who have sex with women (MSW), and women. The base model is the same across subpopulations with parameters varied for probability of testing and treatment. We also estimated the average lifetime QALYs lost per infection in the entire population.

We estimated QALYs lost due to congenital syphilis by developing a separate decision tree. The model estimated the average number of QALYs lost across different possible outcomes per case of congenital syphilis. We included the consequent health losses in mothers and children in each outcome per case of congenital syphilis.

### Disease progression

The Markov cohort model consists of the following main health states in the natural history of syphilis in adults: primary, secondary, early latent, late latent, untreated tertiary, treated (no long-term sequelae), treated with long-term sequelae, neurosyphilis, and death due to syphilis or all other causes (Figure 1). In our model, tertiary syphilis refers to gummas(non-cancerous growth of dead and fiber-like tissue), cardiovascular syphilis, and psychiatric manifestations. We included late neurosyphilis as a separate outcome. Simulation begins at infection and we assumed every infected person progresses to the primary syphilis stage. If untreated, those with primary syphilis are at risk of progressing further to secondary, early and late latent, and tertiary syphilis. Monthly progression rates between primary, secondary, early latent, and late latent syphilis were determined by the duration estimates for each state (4-7). As is conventional, the transitions in the model are exponentially distributed.

**Figure 1.**
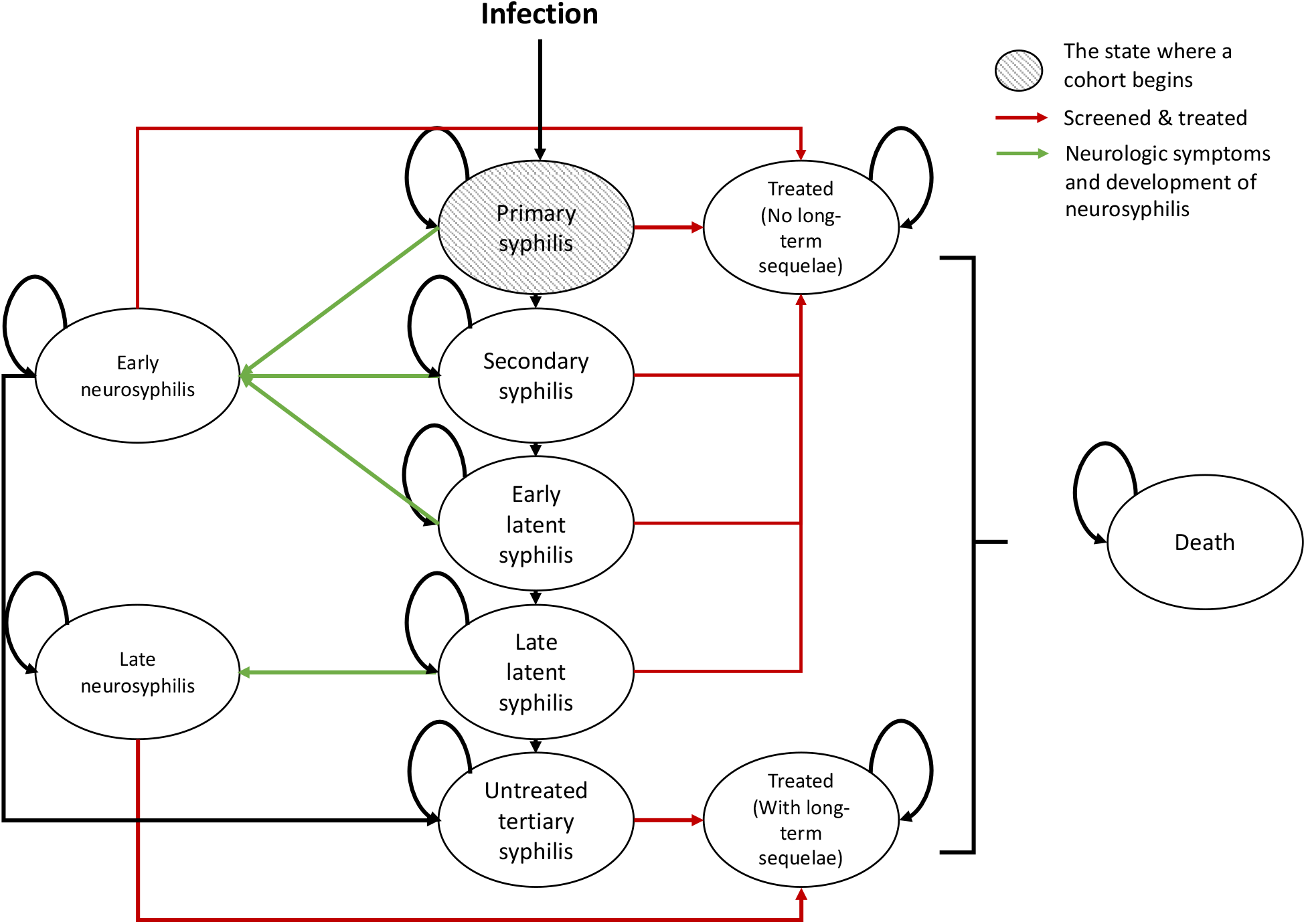
Markov model of syphilis. In our model, tertiary syphilis refers to gummas, cardiovascular syphilis, psychiatric manifestations, etc.; we included late neurosyphilis as a separate outcome. Black arrows indicate health transitions in the natural history of syphilis. Neurologic symptoms and neurosyphilis can develop at any stage of syphilis (green arrows). Treatment can occur at any stage of syphilis (red arrows show the transition to the ‘treated’ state).

In the model, 33% of untreated syphilis has neurologic involvement every month, which is not shown in Figure 1 (11,14,15). In the model, neurological involvement is a precondition before developing neurosyphilis and does not associate with any health loss. Among those with neurologic involvement, the risk of developing neurosyphilis depends on the stage of syphilis: in early syphilis (primary, secondary, and early latent), the monthly risk is 5%. In late latent syphilis, the risk of developing neurosyphilis in 15 years is 9% (i.e. 0.05% monthly risk). Developing neurosyphilis in late syphilis results in long-term health sequelae even if treatment cures infection. In each month, 18% of those in the early neurosyphilis state are treated (8, 9). The rest will remain in early neurosyphilis or progress to tertiary syphilis until the next cycle starts. All states have a background risk of death based on age-specific all-cause mortality in the US (10). We also assumed that people with untreated tertiary syphilis have the same excess all-cause mortality as observed in a study of untreated syphilis (11) (Appendix 1). In order to calculate the number of lifetime QALYs lost attributed to both morbidity and mortality of syphilis, we calculated the quality-adjusted life expectancy for people with syphilis and people without syphilis separately and calculated the difference between the two. We present model input parameters and their distributions in Table 1.

**Table 1.** **Model parameters used to estimate the lifetime number of quality-adjusted life years (QALYs) lost due to syphilis: base case values, distributions applied in sensitivity analyses, and references**

| Variable | Base case value<br>[uncertainty<br>range*] | Distribution type | Reference |
| --- | --- | --- | --- |
| **Natural history** |  |  |  |
| Average duration of infection stage (months) |  |  |  |
| Primary (“dur_p”) | 0.7 [0.25 – 1.96] | Lognormal (-0.36, 0.53) | (4-7) |
| Secondary (“dur_s”) | 3.6 [2.16– 5.99] | Lognormal (1.3, 0.26) | (4-7) |
| Early latent | 7.7 [3 – 11.4] | 12 – dur_p – dur_s | (4-7) |
| Monthly probability of developing neurosyphilis |  |  |  |
| In early syphilis (to early neurosyphilis) | 0.017 | p_neuro*pNS_early | (4, 7, 8) |
| In late syphilis (to late neurosyphilis) | 0.00017 | p_neuro*pNS_late | (4, 6-8) |
| Monthly probability of neurologic involvement (‘p_neuro’) | 0.33 [0.06 – 0.6] | Beta (3.5, 7.1) | (4, 7, 8) |
| Monthly probability of progressing from early syphilis to early neurosyphilis given neurologic involvement (‘pNS_early’) | 0.05 [0.01 – 0.09] | Beta (5.7, 107.4) | (4, 8) |
| Monthly probability of progressing from late syphilis to late neurosyphilis given neurologic involvement (‘pNS_late’) | 0.0005 [0.0003 – 0.0007] | Table S1 | (4) |
| Monthly probability of recovery from early neurosyphilis through treatment | 0.18 [0.12-0.28] | Table S1 | (4, 36) |
| Monthly probability of progressing from early neurosyphilis or late latent to tertiary syphilis | 0.0012 [0.008-0.02] | Table S1 | (4, 6) |
| Monthly probability of treatment failure |  |  |  |
| Primary, secondary, early latent | 0.05 [0 – 0.1] | Beta (3.6, 68.4) | (4) |
| Late syphilis (latent syphilis) | 0.19 [0.08 – 0.3] | Beta (9.1, 38.8) | (4) |
| Mortality |  |  |  |
| All-cause among untreated tertiary syphilis | Table S2** | Fixed | (11) |
| All-cause in other health states | Age- and sex-specific mortality in the US | Fixed | (10) |
| **Disutility of syphilis** |  |  |  |
| Primary (“du_p”) | 0.006 [0.004-0.01] | Beta (3.8, 631.6) | (4, 14) |
| Secondary (“du_s”) | 0.04 [0.02 – 0.06] | Beta (14.7, 343.7) | (4, 14) |
| Early latent (“du_el”) | 0 | Fixed | (4, 14) |
| Late latent | 0 | Fixed | (4, 14) |
| Early neurosyphilis | 0.05 [0.03 – 0.08] | Beta (14.5, 276.4) | (15) |
| Late neurosyphilis | 0.20 [0.12-0.29] | Beta (16.5, 64.6) | (16) |
| Tertiary syphilis (cardiovascular/gummatous) | 0.24 [0.15-0.33] | Beta (21.0, 65.3) | (16) |
| Long-term disutility of tertiary and late neurosyphilis syphilis following treatment | 0.09 [0.05 – 0.14] | Beta (13.8, 133.3) | (4, 14) |
| **Background utility** | Table S3 | Fixed | (21) |
| **Annual rate of testing and treatment** |  |  |  |
| Primary (MSW) | 0.30 [0 - 0.65] | Beta (1.7, 3.9) | (9) |
| Secondary (MSW) | 0.51 [0.16-0.86] | Beta (3.5, 3.4) | (9) |
| Early latent (MSW) | 0.30 [0 -0.65] | Beta (1.7, 3.9) | (9) |
| Late latent (MSW) | 0.19 [0 -0.47] | Beta (1.2, 5.3) | (9) |
| Primary (MSM) | 0.55 [0.1-1] | Beta (2.0, 1.7) | (9) |
| Secondary (MSM) | 0.76 [0.42-1] | Beta (3.8, 1.2) | (9) |
| Early latent (MSM) | 0.44 [0.01-0.87] | Beta (1.8, 2.3) | (9) |
| Late latent (MSM) | 0.44 [0.01-0.87] | Beta (1.8, 2.3) | (9) |
| Primary (Non-pregnant women)*** | 0.30 [0 - 0.65] | Beta (1.7, 3.9) | (9) |
| Secondary (Non-pregnant women) | 0.51 [0.16-0.86] | Beta (3.5, 3.4) | (9) |
| Early latent (Non-pregnant women) | 0.30 [0 - 0.65] | Beta (1.7, 3.9) | (9) |
| Late latent (Non-pregnant women) | 0.19 [0 - 0.47] | Beta (1.2, 5.3) | (9) |
| Primary (Pregnant women)*** | 0.79 [0.46-1] | Beta (3.8, 1.0) | (37, 38) |
| Secondary (Pregnant women) | 0.79 [0.46-1] | Beta (3.8, 1.0) | (37, 38) |
| Early latent (Pregnant women) | 0.79 [0.46-1] | Beta (3.8, 1.0) | (37, 38) |
| Late latent (Pregnant women) | 0.79 [0.46-1] | Beta (3.8, 1.0) | (37, 38) |
| **Percentage of congenital syphilis diagnoses with clinical characteristics of infants (%)** |  |  |  |
| Live-born with signs or symptoms of congenital syphilis | 33.20 [30.4-35.6] | Dirichlet (431, 796.7, 78.3) | (17) |
| Live-born with no signs or symptoms of congenital syphilis | 60.30 [58.3-63.6] | Dirichlet (431, 796.7, 78.3) | (17) |
| Stillborn | 6.50 [4.78-7.33] | Dirichlet (431, 796.7, 78.3) | (17) |
| **Percentage of symptomatic congenital syphilis with infant outcomes (%)** |  |  |  |
| Low birth weight ('pCS_LB') | 27 [24.6-29.5] | Dirichlet (352.6, 953.3) | (18)<sup>§</sup>, expert opinion |
| Other congenital syphilis symptoms | 73 [70.5-75.4] | 1-'pCS_LB' | (18)<sup>§</sup>, expert opinion |
| **Disutility of mothers due to adverse pregnancy outcomes** |  |  |  |
| Stillbirth | 0.08 [0 - 0.2] | Beta (1.5, 17.1) | (20) |
| Congenital syphilis | 0.12 [0 - 0.3] | Beta (1.4, 10.1) | (20) |
| Low birth weight | 0.0001 [0-0.005] | Beta (0.000006, 0.11) | Assumed |
| Duration of disutility in mother who lost a child | 7 months | Fixed | (22) |
| **Disutility of babies due to adverse pregnancy outcomes** |  |  |  |
| Symptoms of congenital syphilis other than low birth weight | 0.26 [0.12 - 0.4] | Beta (9.5, 27.2) | (20) |
| Low birth weight | 0.11 [0.05-0.16] | Beta (12.1, 102.9) | (20) |
| Total number of QALYs lost to the unborn child per instance of stillbirth | 28.50 | Fixed | (10, 21)<br>Calculated, Appendix 2 |
| Total number of undiscounted QALYs lost to the unborn child per instance of stillbirth | 70.00 | Fixed | (10, 21)<br>Calculated, Appendix 2 |
| **State transition cycle** | 1 month | Fixed | Assumed |
| **Annual discount rate** | 0.03 | Fixed | Assumed |

\* the method to set up uncertainty range is provided in Appendix 4
\*\* we applied age-specific hazard rate ratios of mortality with untreated syphilis to the baseline all-cause mortality to calculate all-cause mortality among those with tertiary syphilis. Details on the calculation of the age-specific hazard rate ratios of mortality with untreated syphilis is provided in Appendix 1
\*\*\* See Table S4 for the calculation of average testing and treatment rate among women
<sup>§</sup> The estimate for the proportion of low birth weight among liveborn babies in the reference was adjusted based on the author assumption.

### Testing and treatment

There is no national data on the rate of syphilis testing and treatment. We used the annual rate of testing and treatment by syphilis stage and subpopulation as assumed in a previously published modeling study (9). We summed the annual rate of seeking treatment and the rate of opportunistic screening and treatment to reflect the different routes of getting tested and treatment (e.g., partner notification, background screening, or treatment seeking due to symptoms). As the previous modeling study suggested, we assumed higher background screening rate among MSM than among the other subpopulations and higher treatment rate for symptomatic syphilis (primary, secondary) than asymptomatic syphilis (early and late latent) (9). The ranges adopted reflect the considerable uncertainty in testing and treatment frequency (Table 1). Women in pregnancy have access to testing and treatment for syphilis as part of prenatal care, and we calculated the average probability of testing and treatment between pregnant and non-pregnant women, weighted by the age-specific prevalence of pregnancy (12) (Table S4). Given high test performance achieved by the combination of nontreponemal and treponemal-specific tests, we assumed that sensitivity and specificity of testing is perfect (13). We assumed people treated for syphilis before developing late neurosyphilis or tertiary syphilis would have no additional loss in quality of life. However, we assumed those treated for syphilis after developing late neurosyphilis or tertiary syphilis would have a lifelong residual detriment to quality of life.

### Disutility of syphilis

We calculated the lifetime loss in QALYs by assuming that each state of syphilis in the Markov model was associated with a detriment in quality of life. The magnitude of this detriment, called a “disutility weight”, was obtained from a review of the literature (Table 1) (4, 14-16). For example, in our model a 70-year-old man would have a quality of life (utility) of 0.773 in the absence of a syphilis-related health outcome (Appendix 2). If the man had tertiary syphilis, there would be a detriment of 0.243 to his quality of life (Table 1), and his utility would be 0.773*(1-0.243), or 0.585. For each month spent with tertiary syphilis, he would incur a loss of 0.188 health utility, calculated as 0.773 – 0.585.

The Markov model calculated QALY losses on a monthly basis over the lifetime of the affected individuals. We calculated both discounted and undiscounted QALYs lost. We discounted the future QALYs lost to the time of infection at 3% per year.

### Congenital syphilis

We set a separate decision tree structure to calculate the average QALYs lost associated with congenital syphilis (Figure 2). The decision tree divides a case of congenital syphilis into three pregnancy outcomes: (1) live-born infants with signs or symptoms of congenital syphilis, (2) live-born infants without signs or symptoms of congenital syphilis, and (3) infants who are stillborn or born alive then die (neonatal death). According to the national statistics on congenital syphilis, among the infants whose vital status is known, 33.2%, 60.3%, and 6.0% of congenital syphilis diagnoses were live-born and symptomatic, live-born and asymptomatic, and stillborn, respectively (17). Among infants who were live-born with signs and symptoms of congenital syphilis, 27% have low birth weight and the rest have other symptoms such as condyloma lata, snuffles, syphilitic rash, and hepatosplenomegaly (18). We assumed that live-born infants with and without signs and symptoms of disease are treated with penicillin for the first 10 days of life and do not incur a long-term reduction in quality of life (19). We assumed live-born symptomatic babies would incur reductions in quality of life for one month due to low birth weight and other symptoms, whereas live-born asymptomatic babies would not have any reductions in quality of life (20). We accounted for lifetime health losses among stillborn infants with the total discounted quality-adjusted life expectancy that was calculated based on the all-cause mortality and age-specific utility scores in the US population (10, 21). We also estimated discounted and undiscounted QALYs lost among mothers per case of congenital syphilis. In the model, mothers with babies with congenital syphilis or low birth weight have short-term associated disutility, whereas disutility among mothers due to stillbirth lasts for 7 months (22). Further details on the estimation are in Appendix 2.

**Figure 2.**
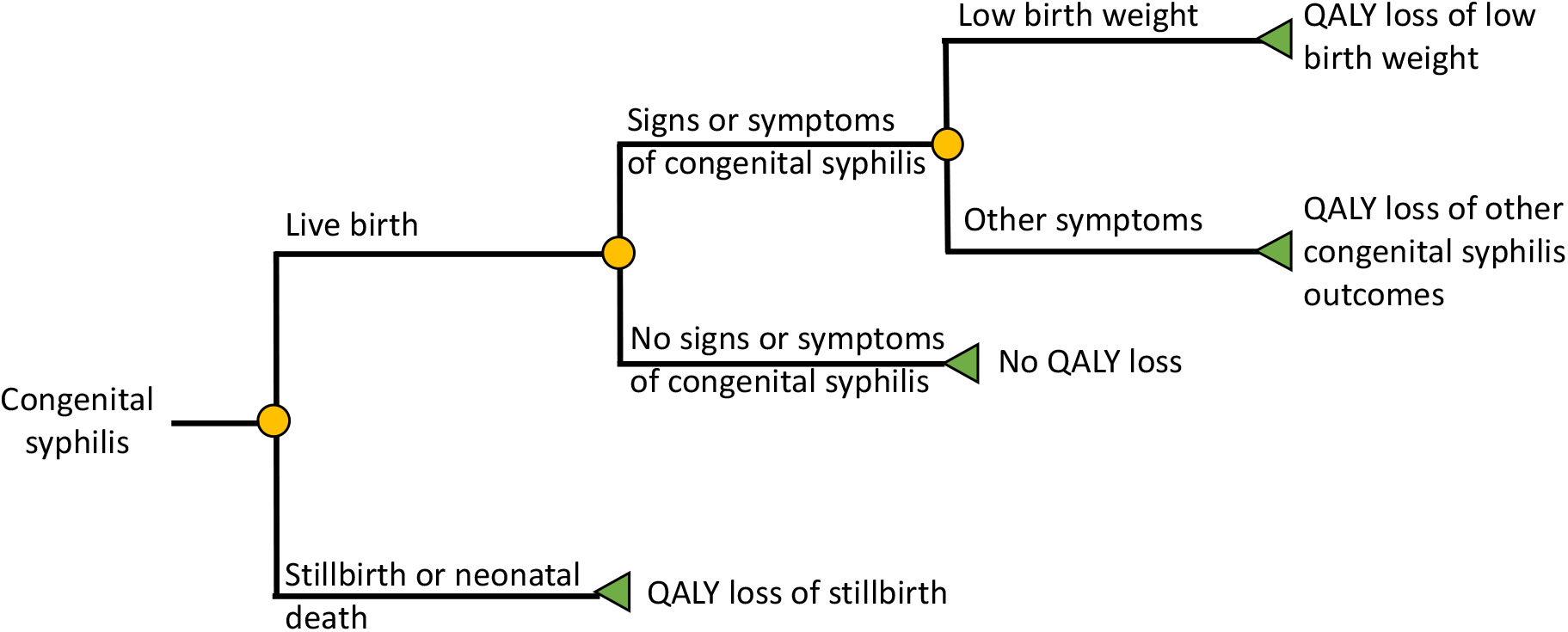
Decision tree model used to estimate the number of quality-adjusted life years (QALYs) lost due to congenital syphilis.

To aggregate the number of QALYs lost due to congenital syphilis, we multiplied the total number of reported congenital syphilis cases in 2018 (1) with the estimated number of QALYs lost in children and mothers per case of congenital syphilis.

### Average number of lifetime QALYs lost per infection in subpopulations and in total population

In each subpopulation, we calculated the average number of QALYs lost across ages at primary infection, by weighting age-specific estimates based on the age distribution of syphilis diagnoses in 2018 (Table S5). We estimated age distribution of syphilis diagnoses based on reported primary and secondary syphilis diagnoses (1, 23). In the male subpopulations (MSM, MSW), we first averaged age-specific numbers of QALYs lost per infection based on the age distribution of syphilis diagnoses among men. We then weighted the estimates between MSM and MSW based on the proportion of syphilis diagnoses in men attributed to MSM (78%) and MSW (22%) (1). The average number of QALYs lost per infection in women does not include the possible QALY losses to mothers and infants due to congenital syphilis. We averaged age-specific QALYs lost per infection based on the age distribution of syphilis among women. In the total population, we averaged the subpopulation-specific QALYs lost per infection based on the proportion of syphilis attributed to each subgroup. We provided details on the weights by age and subgroup used to calculate the average number of lifetime QALYs lost in Appendix 2.

### Total expected number of QALY lost due to syphilis acquired in 2018

We estimated total lifetime number of QALY lost due to syphilis acquired in 2018 by multiplying the estimated number of QALYs lost per infection with the total number of infections in each subpopulation or in the total population. Because the reported primary and secondary syphilis diagnoses are a subset of all new infections, we used the estimated incidence of syphilis in people aged 15-49 years (23). We then extrapolated the incidence for the age outside this range by assuming that the age distribution of the reported primary and secondary syphilis cases is the same as the distribution in the estimated incidence. Our approach assumes that the relative level of underdiagnosis of syphilis is the same across age groups (1, 23). Further details are in Appendix 3.

### Uncertainty analysis

We performed probabilistic sensitivity analysis to generate ranges around the model’s estimates given the significant uncertainty in the model’s input parameters (Table 1). The ranges reflect uncertainty in the natural history parameters, probability of screening and treatment in different subpopulations, and the disutility associated with the adverse outcomes. We sampled parameters 1,000 times independently from pre-defined distributions and ran the model with the sampled parameter sets. The 2.5^th^ and 97.5^th^ percentiles of results from the 1,000 sampled parameter sets are presented as the 95% uncertainty intervals. The distributions used in the sensitivity analysis are described in Appendix 4.

## Results

### Number of QALYs lost per infection by age at primary infection

The number of QALYs lost due to syphilis (per infection) varied by age at infection (Figure 3, Table S6, S7). In all subpopulations, the number of QALYs lost per infection decreased with age at infection. The number of QALYs lost per infection was about 25%-60% lower for MSM than for MSW and women, primarily due to the higher testing and treatment rates in MSM. With primary infection at the ages 20-24 years, for example, the discounted lifetime number of QALYs lost per infection among MSM was 0.07 [0.02-0.21 95%UI], whereas the number of QALYs lost per infection among MSW and women was 0.17 [0.04-0.45 95%UI] and 0.09 [0.04-0.20 95%UI], respectively. Women had lower QALYs lost per infection during the reproductive ages (15-39 years) when pregnant women have increased testing and treatment during pregnancy.

**Figure 3.**
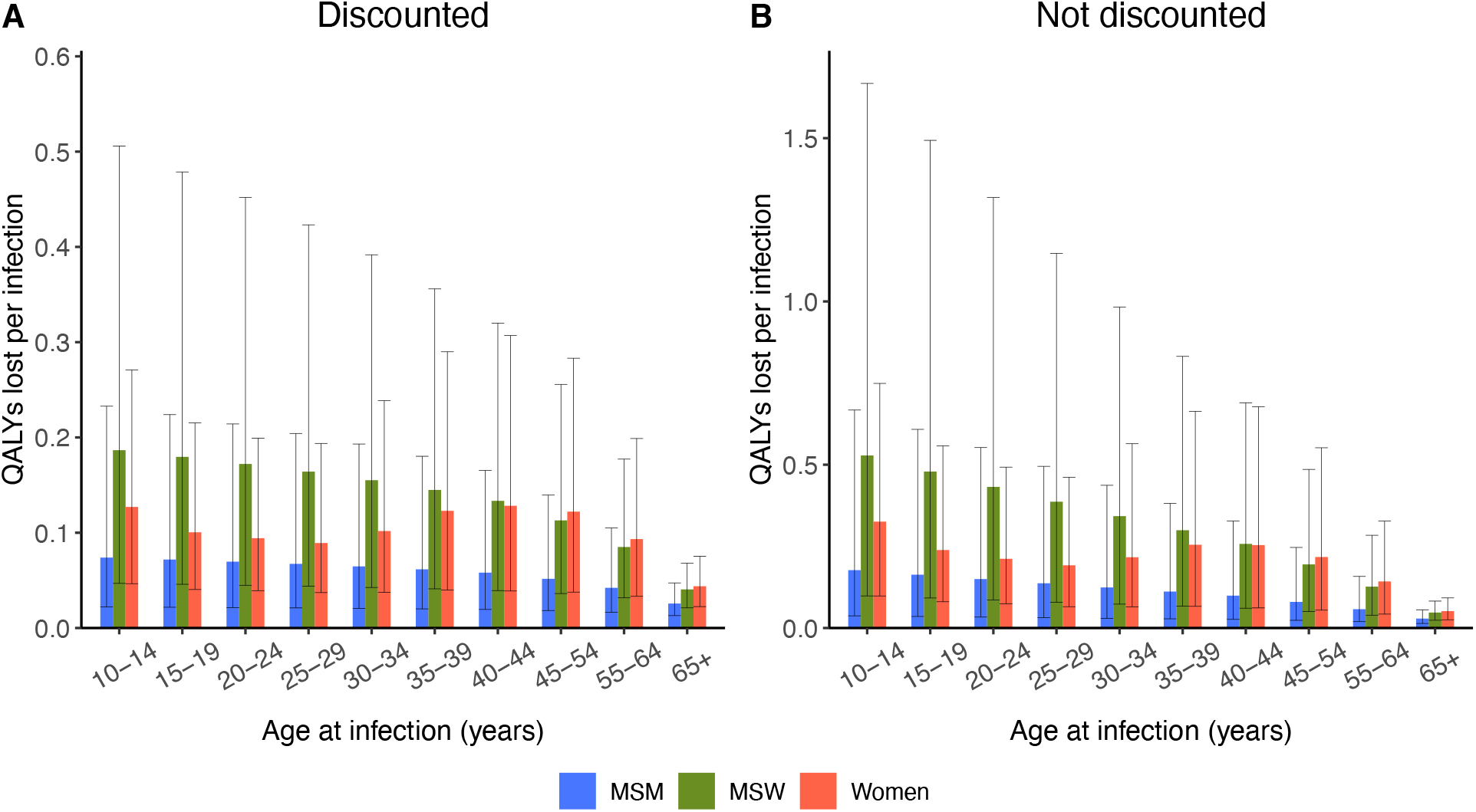
Lifetime number of QALYs lost due to syphilis (per infection) by age at infection for men who have sex with men (MSM), men who have sex with women (MSW), and women: (A) discounted number of QALYs lost per infection; (B) undiscounted number of QALYs lost per infection.

Without discounting, the number of QALYs lost per infection ranged between 0.05 to 0.53. With primary infection at the ages 20-24 years, for example, the undiscounted lifetime number of QALYs lost per infection among MSM was 0.15 [0.03-0.55 95%UI], whereas in MSW and women the estimates were 0.43 [0.09-1.32 95%UI] and 0.21 [0.07-0.49 95%UI], respectively.

### Average lifetime number of QALYs lost per infection via adult subpopulations and total population

The average number of QALYs lost due to syphilis per infection among MSM was lower than in MSW and women (Figure 4A, Table S8). On average, the discounted number of QALYs lost per infection was 0.06 [0.02-0.19 95% UI] in MSM, whereas it was 0.15 [0.04-0.38 95% UI] and 0.10 [0.04-0.23 95% UI] in MSW and women, respectively. In the entire US population, the average discounted number of QALYs lost per infection was 0.09 [0.03-0.19 95% UI]. The undiscounted number of QALYs lost per infection was 0.12 [0.03-0.43 95% UI] in MSM, whereas it was 0.33 [0.07-0.97 95% UI] and 0.22 [0.07-0.54 95% UI] in MSW and women, respectively (Figure 4B, Table S8). In the entire US population, the average undiscounted number of QALYs lost per infection was 0.18 [0.06-0.43 95% UI].

**Figure 4.**
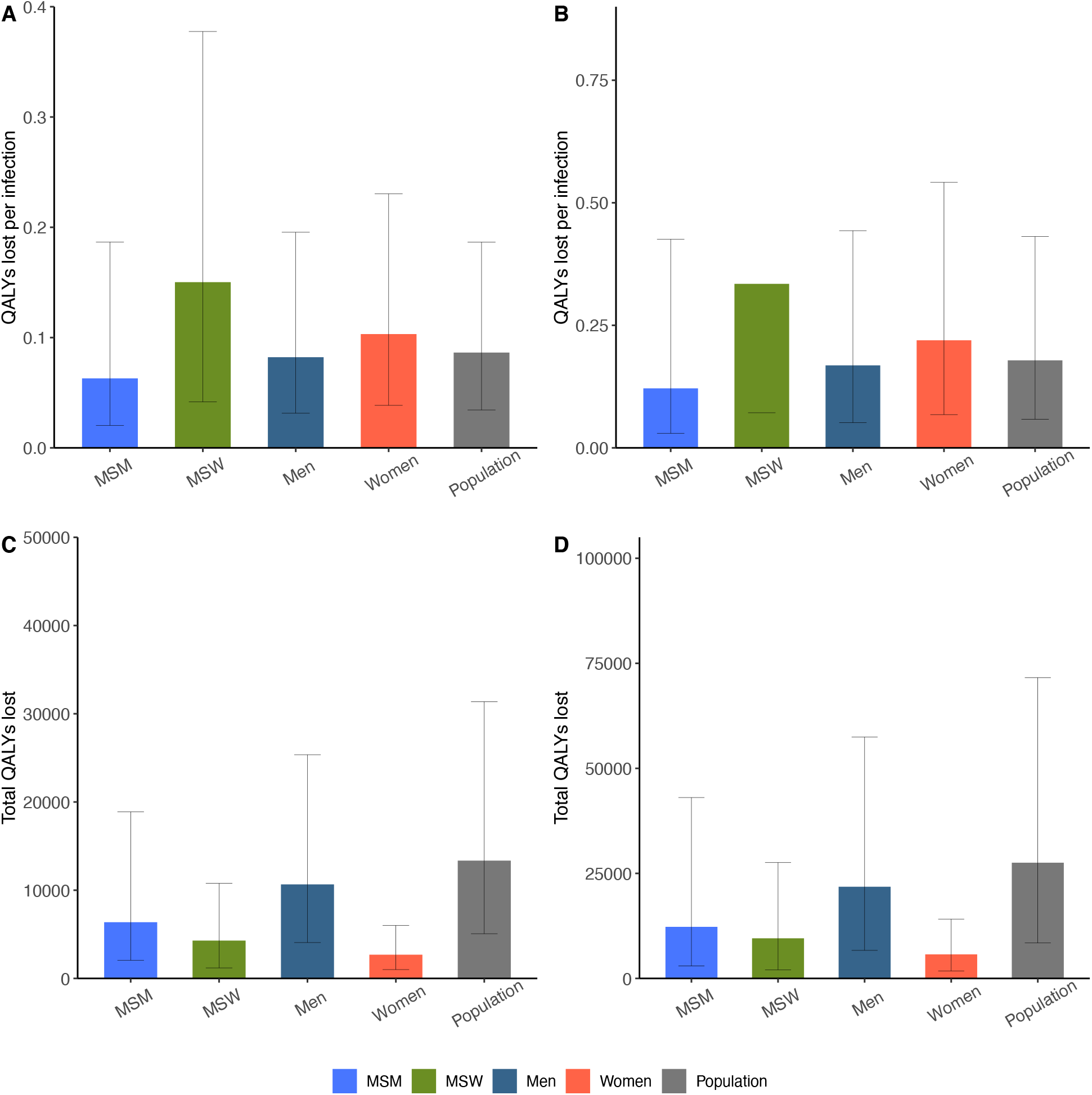
Average lifetime number of QALYs lost due to syphilis (per infection) and total number of QALYs lost due to syphilis acquired in 2018 for men who have sex with men (MSM), men who have sex with women (MSW), women, and total population: (A) discounted QALYs lost per infection (B) undiscounted QALYs per infection (3) discounted total QALYs lost (D) undiscounted total QALYs lost.

### Total lifetime number of QALYs lost due to syphilis acquired in 2018

In total, syphilis incidence in 2018 was expected to result in 13,349 discounted QALYs lost [5,071 – 31,360 95% UI] in the US population (Figure 4C). MSM account for 47.7% of the total number of discounted QALYs lost (6,373 QALYs lost). MSW and women account for 32.1% (4,286 QALYs lost) and 20.2% (2,691 QALYs lost) of the total number of discounted QALYs lost, respectively.

When future health loss was not discounted, the total number of QALYs lost due to syphilis in 2018 was estimated to be 27,544 [8,446-71,594 95%UI] (Figure 4D). The expected total number of QALYs lost due to syphilis in 2018 in MSM, MSW, and women was 12,276, 9,546, and 5,722 respectively. Mean estimate and uncertainty range of the total number of QALYs lost are described in Appendix (Table S9). In total, all syphilis incidence that was sexually and vertically acquired in 2018 is expected to cause 13,349 QALYs lost if the estimate is discounted, and 27,544 QALYs lost if the estimate is not discounted.

Disutility from long-term sequelae with late neurosyphilis or tertiary syphilis accounted for the largest proportion of the estimated total number of QALYs lost (Figure 5). Compared to MSW and women, a lower proportion of the QALY burden in MSM was attributable to late syphilis (untreated tertiary syphilis, late neurosyphilis, and their long-term sequalae). In all subpopulations, the proportion of QALYs lost due to excess mortality with syphilis was relatively low.

**Figure 5.**
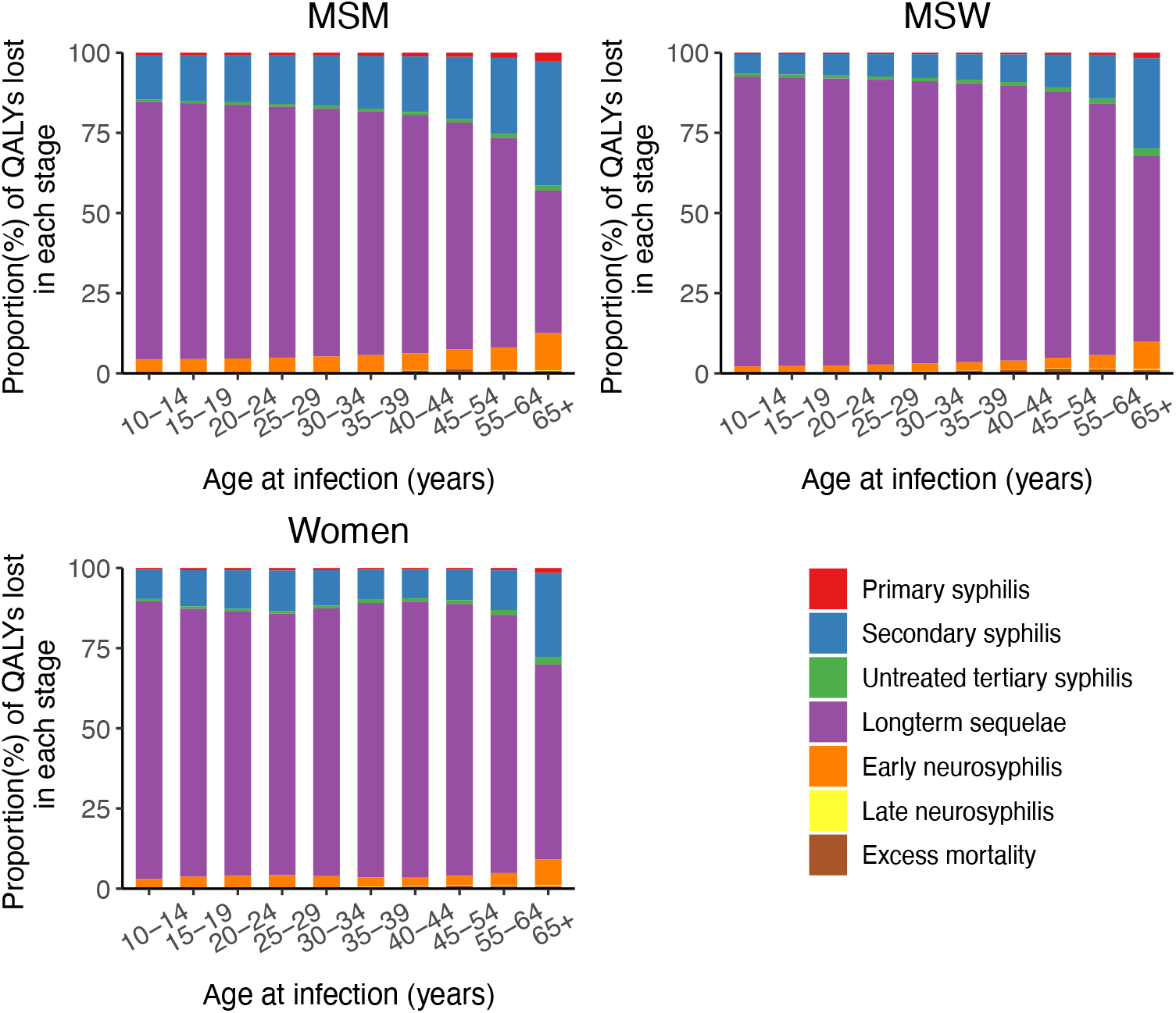
Proportion of the number of QALYs lost attributed to each stage of syphilis for men who have sex with men (MSM), men who have sex with women (MSW), and women.

### QALYs lost due to congenital syphilis

On average, each case of congenital syphilis resulted in a loss of 0.06 QALYs [0.01 – 0.14 95% UI] in mothers and 1.79 QALYs [1.43 – 2.16 95%UI] in the affected child (Figure 6A, Table S10). If future health loss was not discounted, the value would be 0.06 and 4.28 for mothers and children, respectively. The total number of lifetime QALYs lost due to congenital syphilis cases that occurred in 2018 was 79.49 for mothers and 2,332 for children (Figure 6B). Without discounting, the total expected number of QALYs lost for mothers and children is 84 and 5,596, respectively. We provided the number of QALYs lost per infection in women with and without including QALY losses due to congenital syphilis in Table S11.

**Figure 6.**
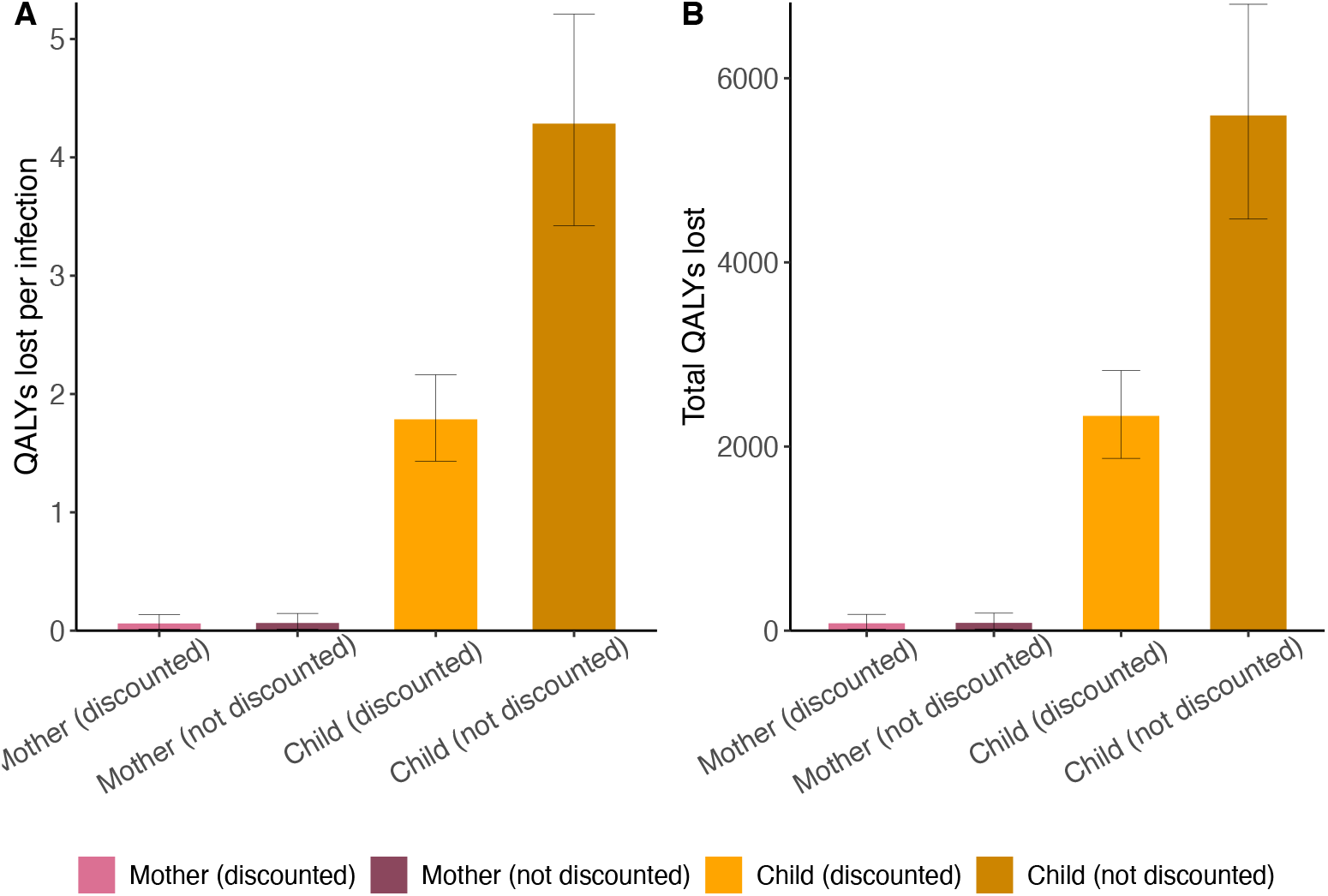
Number of QALYs lost due to congenital syphilis. (A) Average number of QALYs lost per case of congenital syphilis (B) total expected number of QALYs lost due to congenital syphilis cases that occurred in 2018

## Discussion

We quantified the lifetime number of QALYs lost due to syphilis per infection in the US population. Although the average lifetime QALYs lost per infection in MSM was lower than for MSW and women, MSM accounted for the highest total number of QALYs lost. The main reason why the average burden per infection was lower for MSM than for MSW and women was that MSM had a higher rate of treatment as a result of a higher background rate of screening. For example, the CDC guidelines recommend annual syphilis serology test for sexually active MSM, including those with HIV infection (24). Similarly, women had a relatively low average number of QALYs lost per infection during the reproductive age (15-39 years), because pregnant women have increased testing and treatment during pregnancy, thereby increasing the average rate of treatment for this group. Most of the QALY losses were attributed to detriments to quality of life in the stage of living with long-term sequelae after developing late neurosyphilis or tertiary syphilis.

Our results can inform cost-effectiveness analyses of syphilis prevention interventions. Consider a hypothetical syphilis prevention intervention that is estimated to cost $1 million. Suppose the intervention is estimated to avert 500 incident infections, based on a mathematical model of intervention impact or some other method of estimation. Our QALY estimates could be used to generate cost-effectiveness ratios for this intervention (3). For example, our estimate of the average number of QALYs lost per infection (0.09) can be combined with existing estimates of the lifetime medical cost per infection ($1,190), as follows. This intervention would be expected to avert $595 million in lifetime medical costs (500 x $1,190) and to save 45 QALYs (500 × 0.09). The cost per QALY gained by this intervention would be $9,000, calculated as ($1,000,000 - $595,000)/45. We note that previous estimates of the lifetime number of QALYs lost per HIV infection (25) have been used in a similar manner in numerous cost-effectiveness analyses of HIV prevention interventions (26, 27).

Our results can also help to quantify the health burden of syphilis and facilitate comparison to other adverse health outcomes. For example, the average lifetime number of QALYs lost per infection that we estimated (0.09 QALYs) can be compared to 5.8 QALYs lost per HIV infection (25), 0.024 QALYs lost per case of genital warts (28-30) and 0.036 QALYs lost per nonfatal motor vehicle accident (31). Similarly, our estimate of the total number of QALYs lost due to syphilis acquired in 2018 (13,349 QALYs) can be compared to estimates of the total number of QALYs lost due to other infections.

Our study illustrates the challenges associated with estimating the lifetime QALY loss due to syphilis. Most importantly, our models require more complete data than are currently available. Owing to limited data, we used syphilis treatment rates applied in a published modeling study simulating syphilis transmission in two US states (9). We accounted for the uncertainty in treatment rates for syphilis in the sensitivity analyses where wide ranges were used. The wide uncertainty range we applied included the treatment rates estimated in another modeling study (9). Similarly, limited data exist to inform our estimates of the disutility associated with adverse outcomes of syphilis, as well as our estimates of the rates at which these adverse outcomes occur. As with the uncertainty in treatment rates, we accounted for uncertainty in these other model inputs by performing sensitivity analyses. Our analysis can be further updated as better data become available. There is currently no consensus on how to calculate the QALY losses from stillbirths and infant deaths. For example, although some studies have considered lifelong maternal disutility due to losing a child, most studies have excluded parental effects completely (33-35). We took an intermediate approach and included a temporary disutility for mothers who lost a child. Using this intermediate approach, our estimate on lifetime health loss due to congenital syphilis among mothers would be lower than the estimate when lifelong disutility of losing a child is considered and lower than when parental effects are completely excluded. Our study focused on health losses directly associated with sequelae following infection. We did not include other possible health outcomes associated with syphilis, such as new HIV infections attributable to the facilitative effects of syphilis on HIV transmission and acquisition. Consideration of broader health impact of syphilis would increase our estimate of health losses due to syphilis.

In summary, we developed estimates of the quality-of-life impact of adult syphilis and congenital syphilis in the United States. Our estimates of the number of QALYs lost due to syphilis (per infection, and across the total population) can be used in a wide range of health economic studies and burden of disease studies of syphilis. We included uncertainty intervals in our estimates to reflect limitations in currently available data. Finally, we provided detailed documentation of our methods and assumptions so that those who use our estimates in future studies can make updates and modifications as needed in their application of our estimates.

## Data Availability

All data produced are available in literature (reference)

## Supplementary material for the manuscript

**Table S1.** Model parameters used to estimate the lifetime number of quality-adjusted life years (QALYs) lost due to syphilis (extended version): base case values, calculation of monthly probabilities, distributions applied in sensitivity analyses, and references.

| Variable | Base case value<br>[uncertainty<br>range*] | Distribution type | Reference |
| --- | --- | --- | --- |
| **Natural history** |  |  |  |
| Average duration of infection stage (months) |  |  |  |
| Primary ("dur_p") | 0.7 [0.25 – 1.96] | Lognormal (-0.36, 0.53) | (4-7) |
| Secondary ("dur_s") | 3.6 [2.16– 5.99] | Lognormal (1.3, 0.26) | (4-7) |
| Early latent | 7.7 [3 – 11.4] | 12 – dur_p – dur_s | (4-7) |
| Monthly probability of developing neurosyphilis |  |  |  |
| In early syphilis (to early neurosyphilis) | 0.017 | p_neuro*pNS_early | (4, 7, 8) |
| In late syphilis (to late neurosyphilis) | 0.00017 | p_neuro*pNS_late | (4, 6-8) |
| Monthly probability of neurologic involvement ("p_neuro") | 0.33 [0.06 – 0.6] | Beta (3.5, 7.1) | (4, 7, 8) |
| Monthly probability of progressing from early syphilis to early neurosyphilis given neurologic involvement ("pNS_early") | 0.05 [0.01 – 0.09] | Beta (5.7, 107.4) | (4, 8) |
| 15-year probability of progressing from late syphilis to late neurosyphilis given neurologic involvement | 0.09 [0.06 – 0.12] | Beta (31.4, 317.2) | (4) |
| Monthly probability of progressing from late syphilis to late neurosyphilis given neurologic involvement ("pNS_late") | 0.0005 [0.0003 – 0.0007] | 1-exp(ln(1-0.09)/180) | Calculated from 15-year probability |
| 6-month probability of recovery from early neurosyphilis | 0.70 [0.54-0.86] | Beta (21.4, 9.2) | (4, 36) |
| Monthly probability of recovery from early neurosyphilis through treatment | 0.18 [0.12-0.28] | 1-exp(ln(1-0.7)/6) | Calculated from 6-month probability |
| 20-year probability of progressing from early neurosyphilis or late latent to tertiary syphilis | 0.25 [0.15-0.35] | Beta (17.8, 53.3) | (4, 6) |
| Monthly probability of progressing from early neurosyphilis or late latent to tertiary syphilis | 0.0012 [0.008-0.02] | 1-exp(ln(1-0.25)/240) | Calculated from 20-year probability |
| **Probability of treatment failure** |  |  |  |
| Primary, secondary, early latent | 0.05 [0 – 0.1] | Beta (3.6, 68.4) | (4) |
| Late syphilis (latent syphilis) | 0.19 [0.08 – 0.3] | Beta (9.1, 38.8) | (4) |
| **Mortality** |  |  |  |
| All-cause among untreated tertiary syphilis | Table S2** | Fixed | (11) |
| All-cause in other health states | Age- and sex-specific mortality in the US | Fixed | (10) |
| **Disutility of syphilis** |  |  |  |
| Primary ('du_p') | 0.006 [0.004-0.01] | Beta (3.8, 631.6) | (4, 14) |
| Secondary ('du_s') | 0.04 [0.02 – 0.06] | Beta (14.7, 343.7) | (4, 14) |
| Early latent ('du_el') | 0 | Fixed | (4, 14) |
| Late latent | 0 | Fixed | (4, 14) |
| Early neurosyphilis | 0.05 [0.03 – 0.08] | Beta (14.5, 276.4) | (15) |
| Late neurosyphilis | 0.20 [0.12-0.29] | Beta (16.5, 64.6) | (16) |
| Tertiary syphilis (cardiovascular/gummatous) | 0.24 [0.15-0.33] | Beta (21.0, 65.3) | (16) |
| Long-term disutility of tertiary and late neurosyphilis syphilis following treatment | 0.09 [0.05 – 0.14] | Beta (13.8, 133.3) | (4, 14) |
| **Background utility** | Table S3 | Fixed | (21) |
| **Annual rate of testing and treatment** |  |  |  |
| Primary (MSW) | 0.30 [0 - 0.65] | Beta (1.7, 3.9) | (9) |
| Secondary (MSW) | 0.51 [0.16-0.86] | Beta (3.5, 3.4) | (9) |
| Early latent (MSW) | 0.30 [0 -0.65] | Beta (1.7, 3.9) | (9) |
| Late latent (MSW) | 0.19 [0 -0.47] | Beta (1.2, 5.3) | (9) |
| Primary (MSM) | 0.55 [0.1-1] | Beta (2.0, 1.7) | (9) |
| Secondary (MSM) | 0.76 [0.42-1] | Beta (3.8, 1.2) | (9) |
| Early latent (MSM) | 0.44 [0.01-0.87] | Beta (1.8, 2.3) | (9) |
| Late latent (MSM) | 0.44 [0.01-0.87] | Beta (1.8, 2.3) | (9) |
| Primary (Non-pregnant women)*** | 0.30 [0 - 0.65] | Beta (1.7, 3.9) | (9) |
| Secondary (Non-pregnant women) | 0.51 [0.16-0.86] | Beta (3.5, 3.4) | (9) |
| Early latent (Non-pregnant women) | 0.30 [0 -0.65] | Beta (1.7, 3.9) | (9) |
| Late latent (Non-pregnant women) | 0.19 [0 -0.47] | Beta (1.2, 5.3) | (9) |
| Primary (Pregnant women)*** | 0.79 [0.46-1] | Beta (3.8, 1.0) | (37, 38) |
| Secondary (Pregnant women) | 0.79 [0.46-1] | Beta (3.8, 1.0) | (37, 38) |
| Early latent (Pregnant women) | 0.79 [0.46-1] | Beta (3.8, 1.0) | (37, 38) |
| Late latent (Pregnant women) | 0.79 [0.46-1] | Beta (3.8, 1.0) | (37, 38) |
| **Percentage of congenital syphilis diagnoses with clinical characteristics of infants (%)** |  |  |  |
| Live-born with signs or symptoms of congenital syphilis | 33.20 [30.4-35.6] | Dirichlet (431, 796.7, 78.3) | (17) |
| Live-born with no signs or symptoms of congenital syphilis | 60.30 [58.3-63.6] | Dirichlet (431, 796.7, 78.3) | (17) |
| Stillborn | 6.50 [4.78-7.33] | Dirichlet (431, 796.7, 78.3) | (17) |
| **Percentage of symptomatic congenital syphilis with infant outcomes (%)** |  |  |  |
| Low birth weight ('pCS_LB') | 27 [24.6-29.5] | Dirichlet (352.6, 953.3) | <sup>§</sup>(18), expert opinion |
| Other congenital syphilis symptoms | 73 [70.5-75.4] | 1-'pCS_LB' | <sup>§</sup>(18), expert opinion |
| **Disutility of mothers due to adverse pregnancy outcomes** |  |  |  |
| Stillbirth | 0.08 [0 - 0.2] | Beta (1.5, 17.1) | (20) |
| Congenital syphilis | 0.12 [0 - 0.3] | Beta (1.4, 10.1) | (20) |
| Low birth weight | 0.0001 [0-0.005] | Beta (0.000006, 0.11) | Assumed |
| Duration of disutility in mother who lost a child | 7 months | Fixed | (22) |
| **Disutility of babies due to adverse pregnancy outcomes** |  |  |  |
| Symptoms of congenital syphilis other than low birth weight | 0.26 [0.12 - 0.4] | Beta (9.5, 27.2) | (20) |
| Low birth weight | 0.11 [0.05-0.16] | Beta (12.1,102.9) | (20) |
| Total number of discounted QALYs lost to the unborn child per instance of stillbirth | 28.50 | Fixed | (10, 21)<br>Calculated, Appendix 2 |
| Total number of undiscounted QALYs lost to the unborn child per instance of stillbirth | 70.00 | Fixed | (10, 21)<br>Calculated, Appendix 2 |
| **State transition cycle** | 1 month | Fixed | Assumed |
| **Annual discount rate** | 0.03 | Fixed | Assumed |

\* the method to set up uncertainty range is provided in Appendix 4
\*\* we applied age-specific hazard rate ratios of mortality with untreated syphilis to the baseline all-cause mortality to calculate all-cause mortality among those with tertiary syphilis. Details on the calculation of the age-specific hazard rate ratios of mortality with untreated syphilis is provided in Appendix 1
\*\*\* See Table S4 for the calculation of average testing and treatment rate among women
<sup>§</sup> The estimate for the proportion of low birth weight among liveborn babies in the reference was adjusted based on the author assumption.

## Appendix 1.

### Excess all-cause mortality with untreated tertiary and neurosyphilis

Shafer J.K. and colleagues studied the difference in all-cause mortality between untreated syphilitic and nonsyphilitic patients. The study consisted of 408 untreated syphilitic and 192 nonsyphilitic male African-American patients in Macon County, Alabama. Below is the estimate of all-cause mortality by 5-year age group in untreated syphilitic and nonsyphilitic patients reported in the study.

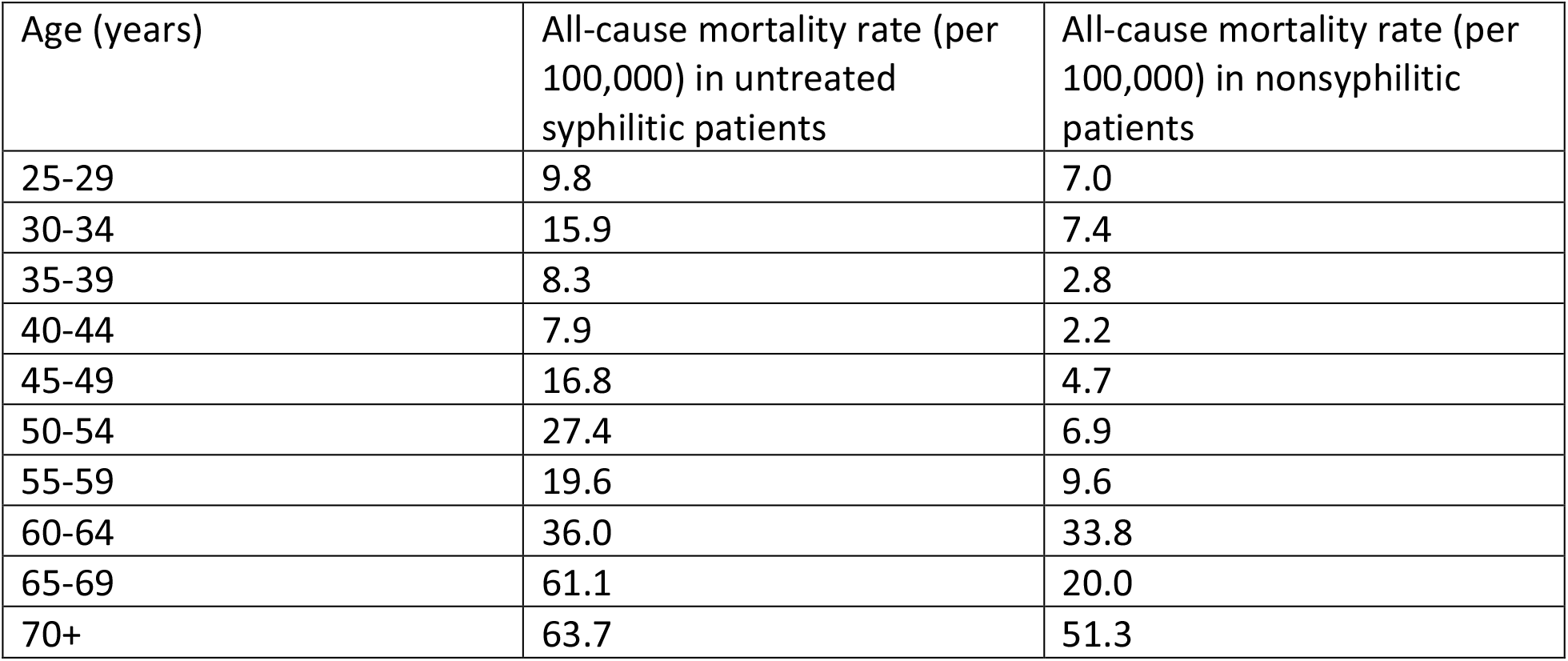

| Age (years) | All-cause mortality rate (per 100,000) in untreated syphilitic patients | All-cause mortality rate (per 100,000) in nonsyphilitic patients |
| --- | --- | --- |
| 25-29 | 9.8 | 7.0 |
| 30-34 | 15.9 | 7.4 |
| 35-39 | 8.3 | 2.8 |
| 40-44 | 7.9 | 2.2 |
| 45-49 | 16.8 | 4.7 |
| 50-54 | 27.4 | 6.9 |
| 55-59 | 19.6 | 9.6 |
| 60-64 | 36.0 | 33.8 |
| 65-69 | 61.1 | 20.0 |
| 70+ | 63.7 | 51.3 |

We then calculated the hazard rate ratio by taking the all-cause mortality between untreated syphilis and nonsyphilitic patients.

**Table S2.** Age-specific hazard rate ratio of mortality with untreated syphilis.

| Age (years) | Rate ratio |
| --- | --- |
| 25-29 | 1.40 |
| 30-34 | 2.15 |
| 35-39 | 2.96 |
| 40-44 | 3.59 |
| 45-49 | 3.57 |
| 50-54 | 3.97 |
| 55-59 | 2.04 |
| 60-64 | 1.07 |
| 65-69 | 3.06 |
| 70+ | 1.24 |

In our model, we applied the age-specific hazard rate ratio of all-cause mortality with untreated tertiary and late neurosyphilis to the background age-specific all-cause mortality in order to calculate all-cause mortality among those with untreated tertiary and late neurosyphilis.

## Appendix 2.

### Total quality-adjusted life expectancy lost due to stillbirths

We calculated the loss of total quality-adjusted life expectancy due to stillbirths or neonatal deaths. Although many health impact studies do not consider stillbirth, omitting it from this analysis would neglect the most impactful adverse outcome of congenital syphilis, and the practical impact of a stillbirth and neonatal death are similar (39). First, we multiplied the probability of surviving over the next year at a given age with the age-specific utility score. We summed the age-specific quality-adjusted life years from age 0 to 100 years. In order to account for 17 days of life before neonatal deaths, we subtracted 17/365 years from the total quality-adjusted life expectancy, when calculating total quality-adjusted life expectancy due to neonatal deaths.

**Table S3.** Age-specific background utility score. Source: Sullivan PW, Ghushchyan V. Preference-Based EQ-5D index scores for chronic conditions in the United States. Med Decis Making. 2006;26(4):410-20

| Age (years) | Male | Female |
| --- | --- | --- |
| 0-19 | 1 | 1 |
| 20-29 | 0.855 | 0.818 |
| 30-39 | 0.839 | 0.807 |
| 40-49 | 0.819 | 0.789 |
| 50-59 | 0.798 | 0.768 |
| 60-69 | 0.792 | 0.772 |
| 70-79 | 0.773 | 0.731 |
| 80-100 | 0.697 | 0.691 |

**Table S4.** Prevalence of pregnancy and the calculation of the average of testing and treatment rate of syphilis among women, as exemplified with testing and treatment in primary syphilis. Source for the prevalence of pregnancy: Curtin SC, Abma JC, Ventura SJ, Henshaw SK. Pregnancy rates for U.S. women continue to drop. NCHS Data Brief. 2013(136):1-8.

| Age (years) | Prevalence of pregnancy (%) | Testing and treatment rate among non-pregnant women | Testing and treatment rate among pregnant women | Average testing and treatment rate among women |
| --- | --- | --- | --- | --- |
| 10-14 | 0.04 | 0.30 | 0.79 | 0.30 |
| 15-19 | 11.03 | 0.30 | 0.79 | 0.35 |
| 20-24 | 11.03 | 0.30 | 0.79 | 0.35 |
| 25-29 | 15.01 | 0.30 | 0.79 | 0.37 |
| 30-34 | 15.01 | 0.30 | 0.79 | 0.37 |
| 35-39 | 4.79 | 0.30 | 0.79 | 0.32 |
| 40-44 | 4.79 | 0.30 | 0.79 | 0.32 |
| 45-54 | 0 | 0.30 | 0.79 | 0.30 |
| 55-64 | 0 | 0.30 | 0.79 | 0.30 |
| 65 and above | 0 | 0.30 | 0.79 | 0.30 |

## Appendix 3.

### Averaging age-specific number of QALYs lost due to syphilis per infection in adult subpopulations

We averaged age-specific number of QALYs lost due to syphilis per infection based on the age distribution of syphilis diagnosis in 2018.

In the male subpopulation, we first calculated the average number of QALYs lost due to syphilis per infection in MSM and MSW, by using the age distribution of syphilis diagnosis among men in 2018. We then combined the estimate in MSM and MSW, by using the proportion of syphilis diagnosis in MSM and MSW among men (78% in MSM and 22% in MSW).

In the female subpopulation, we first calculated the age-specific average number of QALYs lost per infection between pregnant and non-pregnant women. We used prevalence of pregnancy as a weight, assuming that the risk of infections is identical between pregnant and non-pregnant women. We then averaged age-specific estimates in women by using age distribution of syphilis diagnosis in women as a weight.

In calculating the average number of QALYs lost per infection in the population, we weighted subpopulation-specific estimate based on the proportion of total syphilis diagnosis in each subpopulation. In 2018, 64%, 19%, and 17% of total syphilis diagnosis occurred in MSM, MSW, and women, respectively.

**Table S5.** Weights used to calculate average QALYs lost per infection across ages at infection. Source of data: Sexually Transmitted Disease Surveillance 2018. Centers for Disease Control and Prevention; 2019.

| Age (years) | Weight |  |
| --- | --- | --- |
|  | % of syphilis cases* (Male) | % of syphilis cases* (Female) |
| 10-14 | 0.04 | 0.22 |
| 15-19 | 3.63 | 9.22 |
| 20-24 | 14.87 | 21.45 |
| 25-29 | 24.2 | 22.44 |
| 30-34 | 19.90 | 17.91 |
| 35-39 | 14.64 | 13.85 |
| 40-44 | 10.39 | 8.60 |
| 45-54 | 8.00 | 4.57 |
| 55-64 | 3.59 | 1.53 |
| 65 and above | 0.73 | 0.22 |

\* Age distribution was estimated from the reported cases of primary and secondary syphilis.

## Appendix 4.

### Model parameter distributions

We assumed that probability-and utility-type, duration-type, and conditional probability-type of model parameters have beta, lognormal, and Dirichlet distributions, respectively. We also assumed that the range of parameter values identified from literature or multiple data sources represents the 95% confidence interval. If multiple sources of data were not available to set up the range, we set up the range of half to 1.5 times the point estimate as the 95% confidence interval.

**Fig S1.**
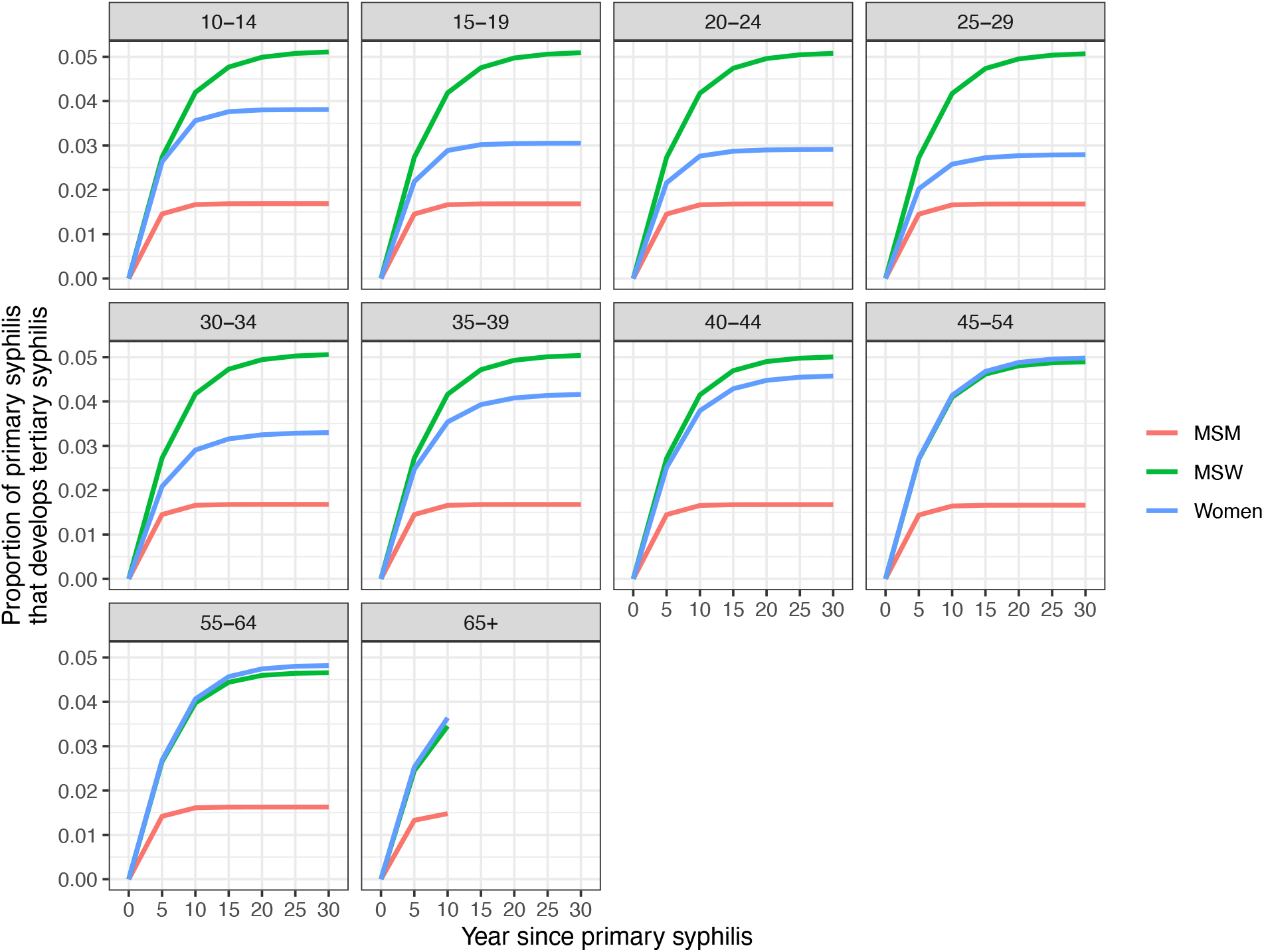
Proportion of primary syphilis that progresses to late neurosyphilis given current testing and treatment coverage.

**Fig S2.**
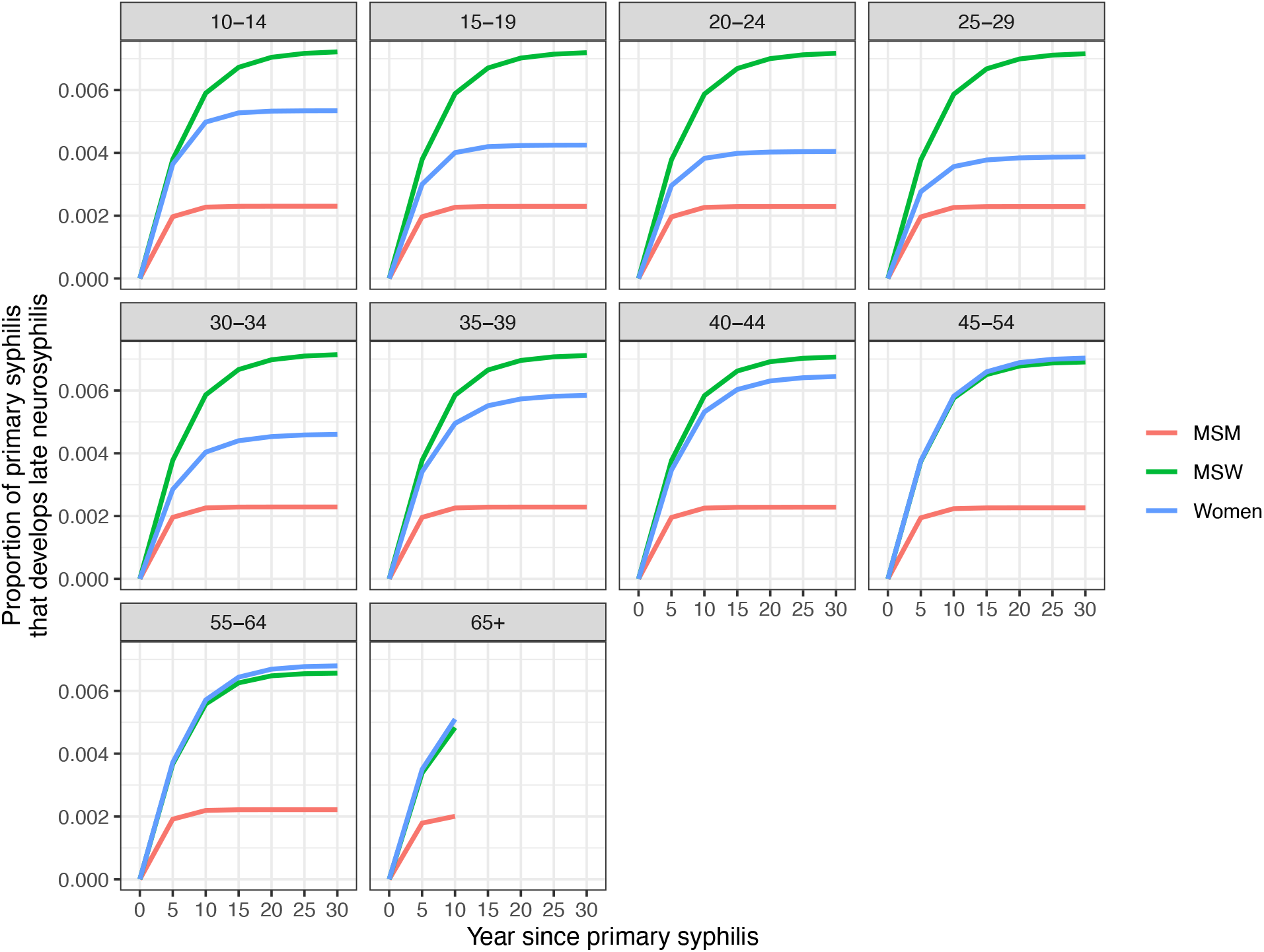
Proportion of primary syphilis that progresses to tertiary syphilis given current testing and treatment coverage.

**Table S6.** Discounted lifetime number of QALYs lost due to syphilis per infection by age at infection.

| Age (years) | MSW | MSM | Women |
| --- | --- | --- | --- |
| 10-14 | 0.19 [0.05-0.51] | 0.07 [0.02-0.23] | 0.13 [0.05-0.27] |
| 15-19 | 0.18 [0.05-0.48] | 0.07 [0.02-0.22] | 0.10 [0.04-0.22] |
| 20-24 | 0.17 [0.04-0.45] | 0.07 [0.02-0.21] | 0.09 [0.04-0.20] |
| 25-29 | 0.16 [0.04-0.42] | 0.07 [0.02-0.20] | 0.09 [0.04-0.19] |
| 30-34 | 0.16 [0.04-0.39] | 0.06 [0.02-0.19] | 0.10 [0.04-0.24] |
| 35-39 | 0.14 [0.04-0.36] | 0.06 [0.02-0.18] | 0.12 [0.04-0.29] |
| 40-44 | 0.13 [0.04-0.32] | 0.06 [0.02-0.17] | 0.13 [0.04-0.31] |
| 45-54 | 0.11 [0.04-0.26] | 0.05 [0.02-0.14] | 0.12 [0.04-0.28] |
| 55-64 | 0.08 [0.03-0.18] | 0.04 [0.02-0.10] | 0.09 [0.03-0.20] |
| 65+ | 0.04 [0.02-0.07] | 0.03 [0.02-0.05] | 0.04 [0.02-0.08] |

**Table S7.** Undiscounted lifetime number of QALYs lost due to syphilis per infection by age at infection.

| Age (years) | MSW | MSM | Women |
| --- | --- | --- | --- |
| 10-14 | 0.53 [0.10-1.67] | 0.18 [0.04-0.67] | 0.33 [0.10-0.75] |
| 15-19 | 0.48 [0.09-1.49] | 0.16 [0.04-0.61] | 0.24 [0.08-0.56] |
| 20-24 | 0.43 [0.09-1.32] | 0.15 [0.03-0.55] | 0.21 [0.07-0.49] |
| 25-29 | 0.39 [0.08-1.15] | 0.14 [0.03-0.50] | 0.19 [0.07-0.46] |
| 30-34 | 0.34 [0.07-0.98] | 0.12 [0.03-0.44] | 0.22 [0.06-0.56] |
| 35-39 | 0.30 [0.07-0.83] | 0.11 [0.03-0.38] | 0.25 [0.07-0.66] |
| 40-44 | 0.26 [0.06-0.69] | 0.10 [0.03-0.33] | 0.25 [0.06-0.68] |
| 45-54 | 0.19 [0.05-0.49] | 0.08 [0.02-0.25] | 0.22 [0.05-0.55] |
| 55-64 | 0.13 [0.04-0.28] | 0.06 [0.02-0.16] | 0.14 [0.04-0.33] |
| 65+ | 0.05 [0.02-0.08] | 0.03 [0.01-0.06] | 0.05 [0.05-0.09] |

**Table S8.**
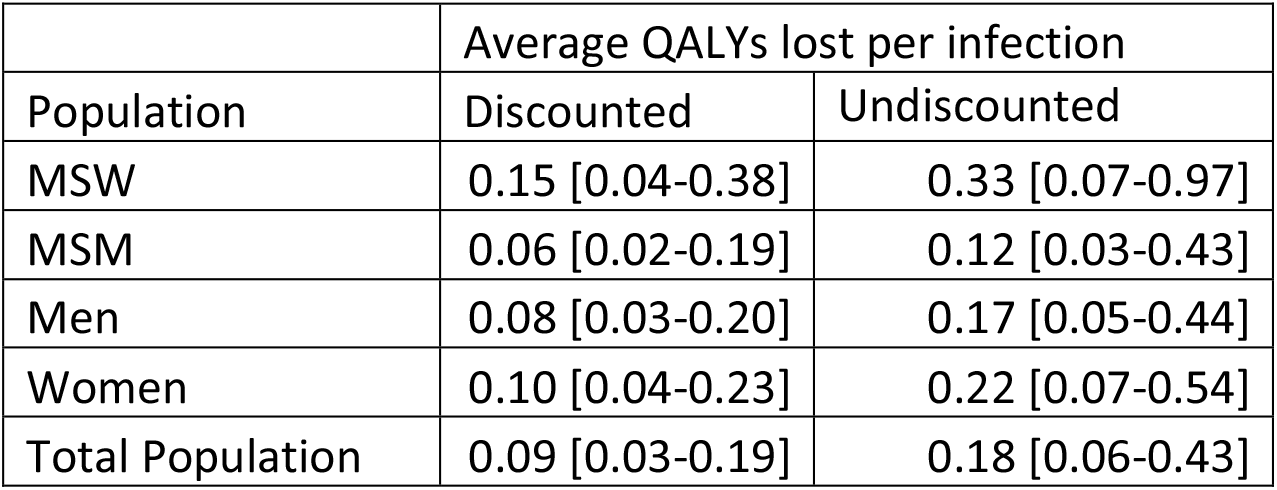
Average lifetime number of QALYs lost due to syphilis per infection by population.

**Table S9.** Total lifetime number of QALYs lost due to syphilis acquired in 2018 by population.

| Population | Total QALYs lost due to infections acquired in 2018 |  |
| --- | --- | --- |
|  | Discounted | Undiscounted |
| MSW | 4,286 [1,191-10,770] | 9,546 [2,042-25,576] |
| MSM | 6,373 [2,062-18,880] | 12,276 [2,980-43,063] |
| Men | 10,659 [4,067-25,354] | 21,822 [6,676-57,463] |
| Women | 2,691 [1,004-6,006] | 5,722 [1,770-14,130] |
| Total Population | 13,349 [5,071-31,360] | 27,544 [8,446-71,594] |

**Table S10.** Number of QALYs lost due to congenital syphilis.

|  | QALYs lost per case of congenital syphilis |  | Total QALYs lost due to congenital syphilis cases acquired in 2018 |  |
| --- | --- | --- | --- | --- |
|  | Discounted | Undiscounted | Discounted | Undiscounted |
| Mothers | 0.06 [0.01-0.14] | 0.06 [0.01-0.15] | 79 [17-177] | 84 [18-190] |
| Children | 1.79 [1.43-2.16] | 4.28 [3.42-5.21] | 2,332 [1,871-2,825] | 5,596 [4,470-6,803] |

**Table S11.** The number of QALYs lost per infection in women with and without including QALY losses due to congenital syphilis. We first calculated the number of QALYs lost due to congenital syphilis (CS) per infection in women by multiplying the number of QALYs lost due to CS per case of CS with the probability of CS given an incidence of syphilis in women. We then added the QALYs losses due to CS per infection in women to the QALYs losses due to syphilis per infection in women based on the prevalence of pregnancy by age as a weight. The number of QALYs lost per case of CS in these calculations included QALYs lost in the mother only, and did not include QALYs lost by the affected child.

| Age (years) | Discounted |  | Undiscounted |  |
| --- | --- | --- | --- | --- |
|  | Without including QALYs lost due to CS | With including QALYs lost due to CS | Without including QALYs lost due to CS | With including QALYs lost due to CS |
| 10-14 | 0.1271<br>[.0465-.2708] | 0.1271<br>[.0465-.2708] | 0.3256<br>[.0982-.7490] | 0.3256<br>[.0982-.7491] |
| 15-19 | 0.1004<br>[.0404-.2154] | 0.1008<br>[.0405-.2161] | 0.2388<br>[.0804-.5580] | 0.2392<br>[.0805-.5589] |
| 20-24 | 0.0942<br>[.0392-.1991] | 0.0946<br>[.0393-.1999] | 0.2118<br>[.0741-.4926] | 0.2121<br>[.0742-.4934] |
| 25-29 | 0.0892<br>[.0372-.1938] | 0.0896<br>[.0373-.1948] | 0.1920<br>[.0653-.4617] | 0.1925<br>[.0654-.4628] |
| 30-34 | 0.1018<br>[.0373-.2385] | 0.1022<br>[.0374-.2395] | 0.2166<br>[.0643-.5642] | 0.2171<br>[.0644-.5653] |
| 35-39 | 0.1230<br>[.0399-.2899] | 0.1231<br>[.0399-.2902] | 0.2549<br>[.0663-.6636] | 0.2550<br>[.0664-.6640] |
| 40-44 | 0.1283<br>[.0389-.3070] | 0.1284<br>[.0389-.3073] | 0.2538<br>[.0616-.6774] | 0.2540<br>[.0616-.6778] |
| 45-54 | 0.1221<br>[.0377-.2832] | 0.1221<br>[.0377-.2832] | 0.2173<br>[.0546-.5519] | 0.2173<br>[.0546-.5519] |
| 55-64 | 0.0933<br>[.0332-.1989] | 0.0933<br>[.0332-.1989] | 0.1425<br>[.0426-.3272] | 0.1425<br>[.0426-.3272] |
| 65+ | 0.0438<br>[.0225-.0752] | 0.0438<br>[.0225-.0752] | 0.0516<br>[.0252-.0921] | 0.0516<br>[.0252-.0921] |
| Average | 0.1032<br>[.0384-.2321] | 0.1035<br>[.0385-.2328] | 0.2194<br>[.0674-.5440] | 0.2197<br>[.0675-.5448] |

